# Characterization of A Novel Autoimmune Encephalitis Associated Antibody against CRMP2

**DOI:** 10.1101/2021.07.15.21260548

**Authors:** Kaibiao Xu, Dongmei Wang, Yan He, Shengnan Wang, Guanghui Liu, Yue Pan, Haishan Jiang, Yu Peng, Fenliang Xiao, Yihua Huang, Qiqi Wang, Yongming Wu, Suyue Pan, Yafang Hu

## Abstract

**Objective:** Autoimmune encephalitis (AE) is a large category disorder urging antibody characterization. The aim was to identify a novel AE related autoantibody targeting an intracellular synaptic protein.

**Methods:** Suspected AE patients had negative conventional antibodies screening but strong immunolabel signals on rat brain sections with the serum and cerebrospinal fluid (CSF) samples were considered burdening unkown antibody. Immunoprecipitation from the rat brain protein lysate followed by mass spectrometry analysis was used to identify the targeting antigen. Western blotting and/or cell-based assay (CBA) with antigen-overexpressing HEK293T cells were used for antibody specificity, epitope and IgG subtype determination. Patients with similar immunostaining pattern on rat brain sections were retrospectively screened for the antibody.

**Results:** The antibody against collapsin response mediator protein 2 (CRMP2), a synaptic protein involved in axon guidance, was identified in a patient with suspected AE. The patient samples reactivated with HEK293T cell overexpressed CRMP2, rather than CRMP1, 3, 4, and 5. The patient samples mainly stained neuronal cytoplasm of the cortex, hippocampus and cerebellum Purkinje cells. This reactivity was eliminated by pre-immunoabsorption with CRMP2-overexpressing HEK293T cells. CRMP2 truncation experiments indicated that 536 amino acids at C-terminus was necessary for the epitope. Subtype analysis showed that anti-CRMP2 antibody was IgG4. Moreover, screening from 19 suspected AE patients led to identification of anti-CRMP2 antibody in another patient with a diagnosis of encephalomyelitis. The two patients responded to immunotherapy.

**Conclusions:** This study discovered a novel anti-CRMP2 antibody associated with AE. Testing of the antibody might be promising for AE diagnosis and treatment.

## INTRODUCTION

Autoimmune encephalitis (AE) is a large category of inflammatory disorders mediated by immune responses against neuronal intracellular antigens, cell-surface or synaptic antigens, in some cases, accompanied by neoplasia. Intracellular antigens include Hu (anti-neuronal nuclear antibody type 1, ANNA1), Ri (ANNA2), and Ma2 in the nucleus, Yo (Purkinje cell cytoplasmic antibody type 1, PCA1), amphiphysin, glutamate decarboxylase 65 kDa isoform (GAD65), and Kelch-like Protein 11 in the cytosol, which cause T cell mediated immune response and some cases respond to immunotherapy ^1-4^. Most AE caused by autoantibodies against antigens on the neuronal surface or synaptic proteins, such as N-methyl-D-aspartate receptor (NMDAR), leucine-rich gliomainactivated 1 (LGI1), contactin-associated protein 2 (Caspr2), gamma-aminobutyric acid (GABA) receptors (A/B), alpha-amino-3-hydroxy-5-methylisoxazole-4-propionic acid (AMPA) receptors, dipeptidyl-peptidase-like protein-6 (DPPX), delta/notch-like epidermal growth factor-related receptor (DNER), dopamine-2 receptor (D2R), metabotropic glutamate receptor 5 (mGluR5), voltage-gated calcium channel salpha-2/delta subunit (Ca_V_α2δ), and glutamate kainate receptor subunit 2 (GluK2), respond well to immunotherapy^1, 5-9^. Thus, intensive testing of these antibodies in suspected AE patients plays an important role in guiding the diagnosis and treatment of the disease. Despite this, the cause of many cases of AE remains unexplained because of limited known antibodies^10-12^. Searching for additional antibodies has been desperately needed.

Family of collapsin response mediator proteins (CRMPs) consists five homologous members, CRMP1-5, which are cytosolic microtubule-associated phosphoproteins and play important roles in dendrite and axonal guidance, regulating migration and synaptic dynamics ^13^. Structurally, CRMP1-4 share 69-76% amino acids (aa) identity, while they share less identity to CRMP5, approximately 50% ^14^. Antibodies against CRMPs except CRMP2 have been reported in several encephalitis. Previously, eight cases of anti-CV2/CRMP5 sera from patients with autoimmune myelopathy were detected anti-CRMP3 antibody positive, and one of them also had anti-CRMP1 antibody, one had anti-CRMP1, 4 antibodies^14-15^. Antibodies to CRMP3-4 were found in a case with subacute limbic encephalitis and thymoma^16^.

CRMPs antibodies also have been found in other disorders. Anti-CRMP1/2 antibodies are found in autoimmune retinopathy patients^17^. Antibodies to CRMP1/2 in maternal sera increased the occurrence of autism spectrum disorders in children^18^. Presence of antibodies to CRMP2 and/or GFAP in the acute phase of spinal cord injury increased the occurrence of neuropathic pain six months later ^19^. Here, we found CRMP2 as the specific autoantigen in a patient with intracranial *Mycoplasma pneumoniae* (*M*.*P*.) infection and possible secondary immune-mediated encephalitis.

## MATERIALS AND METHODS

### Patients and autoantibody testing

This study was approved by the Ethical Review Committee of Nanfang hospital, the Southern Medical University, Guangzhou, China (No. NFEC-2021-001). All patients or their families signed the informed consents.

Patients with suspected autoimmune encephalitis were hospitalized in our Neurology department between January 1, 2018 and December 31, 2020. Patients’ sera or CSF samples were screened for known autoantibodies, including anti-NMDAR, LGI1, Caspr2, GABA_B_R, AMPA_A/B_R, DPPX, DNER, D2R, mGluR5, GAD65 and IgLON5 antibodies by cell-based assay (CBA), some patients tested anti-Hu, Yo, Ri, Ma2, PCA2, amphiphysin, and CRMP5 antibodies by blot assay according to reference^1^. The negative samples were further analyzed by tissue-based assay (TBA) of immunostaining on rat brain sections. CBA negative and TBA positive samples were considered with unknown antibodies and applied to antibody discovery process.

### Immunohistochemistry (IHC)

5μm thick frozen tissue sections from the brain of adult SD rat or C57BL/6 mouse were prepared and performed IHC (or TBA) with CSF, sera, and secondary antibodies as previously reported^20^. Dilutions were used as CSF, 1:1, sera, 1:200, HRP-labeled goat anti-human IgG secondary antibody (bs-0297G-HRP, Bioss, Beijing, China), 1:3000, DyLight 488-labeled goat anti-human IgG secondary antibody (ab97003, Abcam, Cambridge, MA), 1:200. Immunostaining images were photographed under DM3000 microscope (Leica, Wetzlar, Germany) or LSM980 confocal microscope (Zeiss, Oberkochen, Germany). For the absorption assay, the serum diluted 1:200 was pre-immunoabsorbed six times by incubated with acetone fixed CRMP2-overexpressing HEK293T cells for 1 hour at 37□ as reported^6^.

### Western blotting, immunoprecipitation (IP) and liquid chromatography tandem mass spectrometry (LC-MS/MS)

Fresh whole brain protein lysate of adult SD rats and Western blotting were conducted as reported ^20^. The lysis buffer (20 mmol/l Tris-HCl, pH 7.4, 150 mmol/l, 1% TritonX-100, 2 mmol/l EDTA, 5% glyserol, 1 mmol/l PMSF) was used. The protein lysate was mixed with the patient’s CSF at 4□ overnight, followed by adding protein A agarose beads and incubation for another 2 hours. The beads were washed and collected by centrifugation, then boiled with SDS-PAGE sample buffer. The antibody-protein complexes pulled down were separated by 10% SDS gel electrophoresis, transferred to nitrocellulose membrane for Western blotting or stained with Coomassie brilliant blue. Correlated band compared with Western blotting results from the gels was cut and sent for protein identification using LC-MS/MS analysis by Fitgene Biotech Ltd. (Guangzhou, China).

### Cell culture and immunofluorescence (IF)

Primary cortical neurons were prepared from 18 to 19 days C57BL/6 mouse fetuses and cultured for 14 days as reported ^21^. Ice acetone fixed neurons were blocked with 10% goat serum at 37°C for 30 minutes and incubated with patient’ serum (1:200), rabbit anti-MAP2 antibody (1:200, 8707T, Cell Signaling Technology, Danvers, MA) or rabbit anti-CRMP2 antibody (1:200, ab129082, Abcam) at 4°C overnight. DyLight 488-labeled goat anti-human IgG and Alexa Fluor 594-labeled goat anti-rabbit IgG (ab150092, Abcam) secondary antibodies (1:200) were used and fluorescence images were photographed under LSM980 confocal microscope (Zeiss).

HEK293T cells were transfected with plasmids pcDNA3.1-CRMP1/2/3/4-Flag, pcDNA3.1-CRMP5-eGFP, pcDNA3.1-CRMP2 T1 (1-141 amino acids of isoform 1) / T2 (1-36 amino acids of isoform 2)-eGFP, and pcDNA3.1-CRMP2 T3 (142-677 amino acids of isoform 1)-Flag separately for 48 hours. Protein coding sequences in the plasmids referred to mRNA of CRMP1: NM_001014809.3, CRMP2 (isoform 1): NM_001197293.3, CRMP2 (isoform 2): NM_001386.6, CRMP2 (isoform 3): NM_001244604.2, CRMP3: NM_006426.3, CRMP4: NM_001197294.2, and CRMP5: NM_020134.4. Full length isoform 1 of CRMP2 was used unless illustrated. Transfected cells were fixed with ice acetone for 10 minutes, washed with PBST, and blocked with 10% goat serum, then incubated with patient’s serum or rabbit anti-CRMP2 antibody (1:200) overnight at 4°C. After wash, the appropriate secondary antibodies were incubated at room temperature for 1 hour, including DyLight 488-labeled goat anti-human IgG, DyLight 550-labeled goat anti-human IgG (ab96908, Abcam), FITC-labeled anti-human IgG1/2/3/4 (F0767/F4516/F4641/F9890, Sigma-Aldrich) and/or Alexa Fluor 594-labeled goat anti-rabbit IgG secondary antibodies (1:200). The fluorescence images were taken by using LSM980 confocal microscope (Zeiss) or IX73 inverted microscope (Olympus, Tokyo, Japan).

## RESULTS

### Identification of an autoantibody specific targeting CRMP2 in a patient with suspected AE

The index patient (P1) was diagnosed as intracranial *M*.*P*. infection and possible secondary immune-mediated encephalitis. The known autoantibodies tests were negative. However, IHC of P1’s serum showed positive staining across the brain (Figure 1A). Immunofluorescence assay revealed neuronal staining in cortex, hippocampus, and especially Purkinje cells of cerebellum (Figure 1B). Further analysis of P1’s serum showed reaction with the cytosol compartments of cultured cortical neurons (Figure 1C).

**Figure 1.**
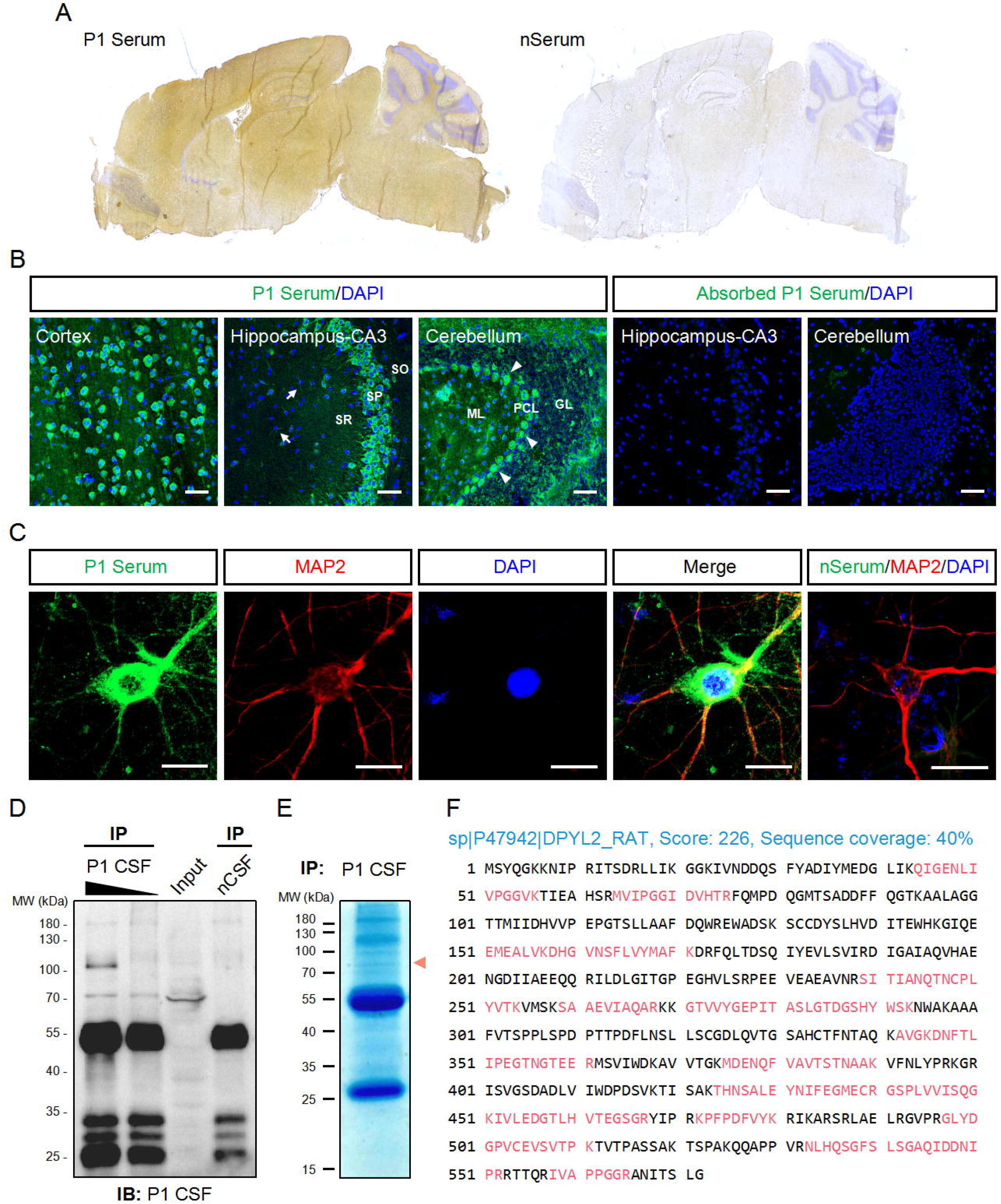
Identification of the autoantibody in a patient with an acute encephalitis. P1 patient was diagnosed as brain *M.P*. infection and possible secondary immune-mediated acute encephalitis. The CSF sample from control patient with Carpal Tunnel Syndrome (nCSF) and the serum from a health person (nSerum) were negative controls. (A) IHC on mouse brain sections. Serum from P1 patient had stained on cortex, hippocampus, and cerebellum. Cell nuclei were counterstained blue with hematoxylin. (B) P1 patient’s serum stained obviously in cytosolic compartments of the neurons in cortex, CA3 area of hippocampus and cerebellum on mouse brain sections (green). White arrows, possible oligodendrocytes; arrow heads, Purkinje cells. CRMP2-expressing HEK293T cell absorbed P1 patient’s serum had no obvious staining. Cell nuclei were stained blue with DAPI. (C) P1patient’s serum (green) stained positively in soma and dendrites of cultured mouse cortical neurons which were labeled with anti-MAP2 Ab (red). (D, E) IP of P1 CSF with rat brain protein lysate. 500 μl (lane 1) and 100 μl (lane 2) P1 CSF were applied to IP, respectively. 500 μl nCSF was used for negative control. A positive band around 70 kDa was pulled down by P1 CSF but not by nCSF. Similar protein size band (red arrow head) was observed on a parallel gel with Coomassie brilliant blue staining (E). (F) Characterization of the autoantigen by mass spectrometry. Suspected protein band in E was collected for protein identification by LC-MS/MS. 17 identified peptides (red) matched to rat DPYL2 (UniProtKB ID: P47942), an alias of CRMP2, with a score 226 and sequence coverage 40%. CRMP2, collapsin response mediator protein 2; CSF, cerebrospinal fluid; DAPI, 6-diamidino-2-phenylindole; DPYL2, dihydropyrimidinase like 2; GL, granular layer; IB, immunoblotting; IHC, immunohistochemistry; IP, immunoprecipitation; LC-MS/MS, liquid chromatography tandem mass spectrometry; *M.P*., *Mycoplasma pneumonia*; MAP2, microtubule associated protein 2; ML, molecular layer; PCL, Purkinje cell layer; SO, stratum oriens; SP, stratum pyramidale; SR, stratum radiatum. Scale bars represent 50 μm in B and 20 μm in C.

Western blotting after IP revealed a band around 70 kDa, which was pulled down by P1’s CSF from rat brain protein lysate (Figure 1D). The correlated visible band from the Coomassie brilliant blue staining gel (Figure 1E) was elucidated and CRMP2 was identified as the possible target antigen by mass spectrometry (Figure 1F, Table S1 in the supplement).

### Verification of the anti-CRMP2 antibody in suspected AE patients

Next, we confirmed that both P1’s serum and commercial anti-CRMP2 antibody detected a band near 70 kDa from the pulled-down protein complex by the P1’s serum from rat brain protein lysate (Figure 2A). With the CRMP2-overexpressing HEK293T cells, CRMP2 were pulled down by commercial anti-CRMP2 antibody and were recognized by P1’s serum (Figure 2B). There are three isoforms of CRMP2, isoform 1 (677 amino acids, 74kDa), isoform 2 (572 amino acids, 62kDa), and isoform 3 (536 amino acids, 58kDa), attribute to three transcript variants, and the observed molecular weight of CRMP2 bands in Western blotting might vary or be higher because of post-translational modification. Here we used the human full-length isoform 1 CRMP2 for further tests unless illustrated.

**Figure 2.**
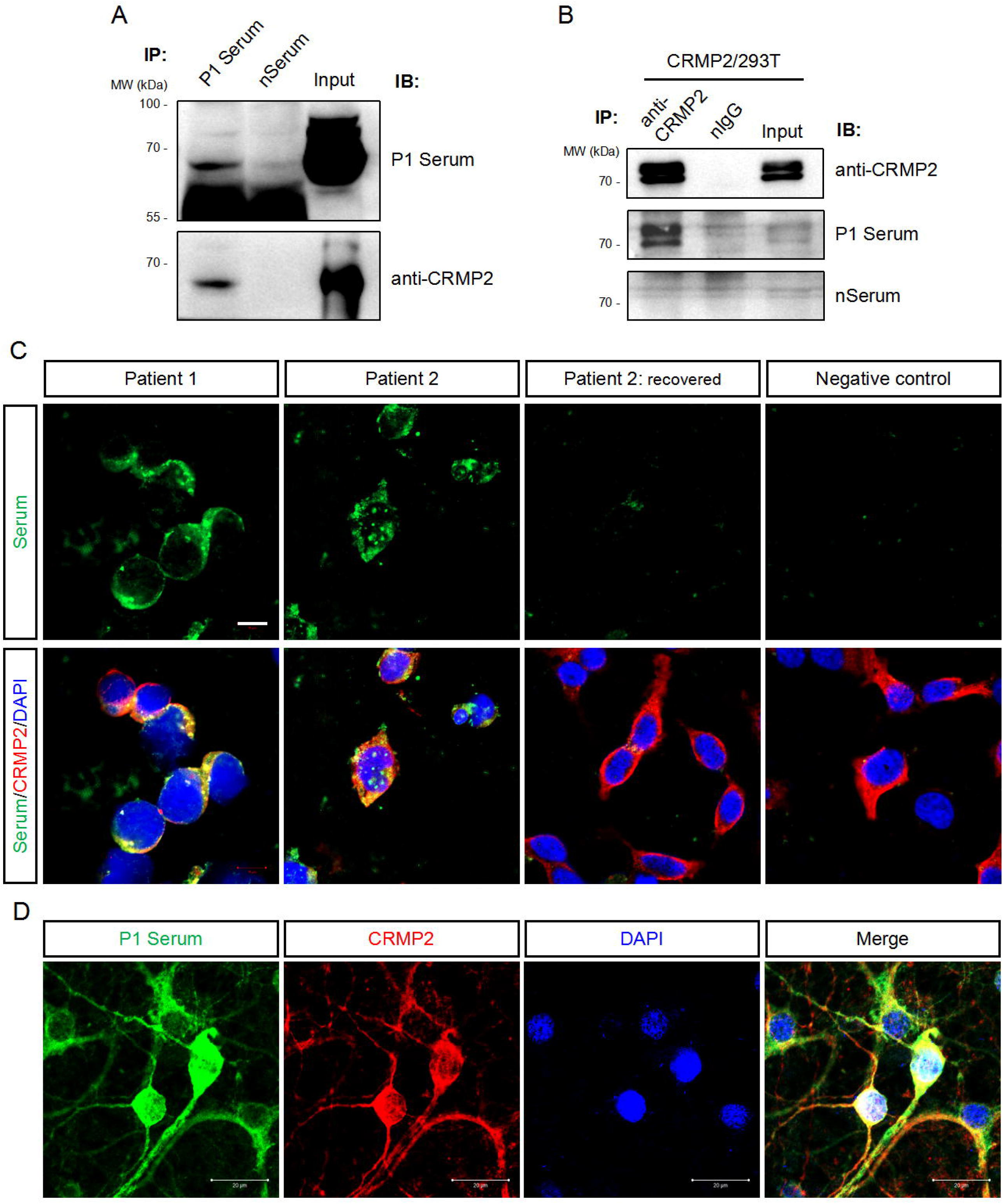
Verification of anti-CRMP2 antibody in the suspected AE patients. (A) IP with P1 patient’s serum was performed with rat brain protein lysate. Immunoblottings were done with P1 patient’s serum (upper panel) and commercial anti-CRMP2 antibody (lower panel). (B) IP with commercial anti-CRMP2 antibody was performed with HEK293T cells overexpressed full length human CRMP2 (isoform 1). Anti-CRMP2 antibody, P1 patient or negative control sera were used for Western blotting, separately. (C) CRMP2-expressing HEK293T cells were immunostained with patients’ sera and anti-CRMP2 antibody. The sera from the first collection of P1 and P2 patients showed positive staining and co-localized with CRMP2. Serum collected from P2 patient after recovered 1.5 years showed negative staining. (D) Colocalization of P1 patient’s serum and anti-CRMP2 Ab immunostaining in cultured mouse cortical neurons. CRMP2, collapsin response mediator protein 2; DAPI, 6-diamidino-2-phenylindole; IB, immunoblotting; nIgG, normal IgG; IP, immunoprecipitation; WB, Western blotting. Scale bars represent 10 μm in C and 20 μm in D.

Anti-CRMP2 antibody and P1’s serum immunostained the overexpressed CRMP2 in HEK293T cells and had well co-localization (Figure 2C). As expected, immunostaining of cultured neurons with P1’s serum also co-localized with anti-CRMP2 antibody signals (Figure 2D). When P1’s serum was pre-immunoabsorbed with the CRMP2-overexpressing HEK293T cells to rule out anti-CRMP2 antibody, a negative immunoreaction was found on the mouse brain section. It indicates that no other neuronal antibodies presented (Figure 1B).

Further, 19 suspected AE patients with negative known antibodies testing but similar immunostaining on rat brain sections were screened for anti-CRMP2 antibody by the CBA method. As indicated in Figure 2C, another patient, P2, was found having immunoreaction with CRMP2 in the serum. Moreover, the immunoreaction was disappeared in P2’s serum collected when the patient recovered completely and was reevaluated 1.5 years later. All the data verify that anti-CRMP2 antibody present in certain AE patients.

### The antibody is IgG4 and specific to the C-terminus of CRMP2

Since CRMP2 belongs to the highly conserved CRMPs family now with five members (Figure 3A), we further determined whether the antibodies in our patients are specific to CRMP2. Expression plasmids were constructed to express full-length CRMP1-5. P1’ and P2’ sera showed immunoreaction only in the HEK293T cells expression of CRMP2, rather than other CRMPs (Figure 3B).

**Figure 3.**
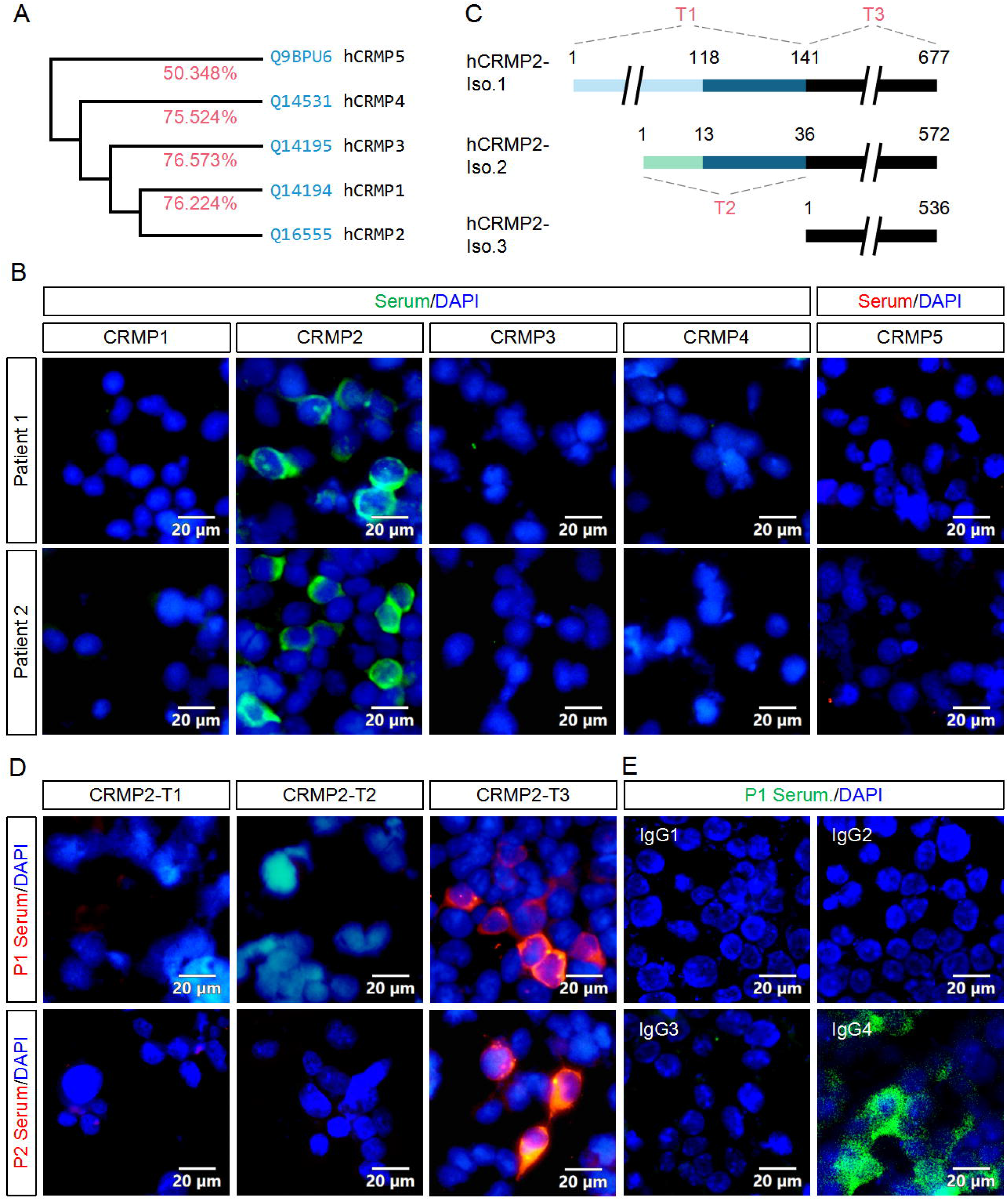
Specificity, epitope and IgG subtype determination of the antibody. (A) Homology analysis showed that CRMP2 had an identity (red) of 76.224%, 76.573%, 75.524% and 50.348% with CRMP1, 3, 4, and 5, respectively. Blue, UniProtKB accession number. (B) CBA assay of the patients’ sera for CRMPs. pcDNA3.1 based plasmids carrying CRMP1, 2, 3, 4 and CRMP5-eGFP were transfected into HEK293T cells separately for protein overexpression. Secondary antibodies anti-human IgG were labeled with green (CRMP1-4) or red (CRMP5) fluorescence. Cell nuclei were stained blue with DAPI. P1 (upper panels) and P2 (lower panels) patients’ sera were only immunoreactive with CRMP2 among CRMP family members. (C) Schematic draws of the three CRMP2 isoforms and of the truncation strategy. The length or amino acid sites were indicated with numerals. Identical sequences between the isoforms were illustrated with same line colors. (D) pcDNA3.1 based plasmids carrying CRMP2 truncates T1, T2 or T3 illustrated in C were transfected into HEK293T cells separately for CBA assay. P1 (upper panels) and P2 (lower panels) patient’s sera were reactive with T3 or 536 amino acids C-terminus of CRMP2 (red). (E) Immunostaining of CRMP2-expressing HEK293T cells with P1 patient serum and FITC conjugated anti-human IgG1-4 secondary antibodies. Positive stained in IgG4. CRMP, collapsin response mediator protein; DAPI, 6-diamidino-2-phenylindole; Iso., isoform. Scale bars represent 20 μm in B, D and E.

To further determine which part of CRMP2 responsible for the antigen-antibody reaction, three expression plasmids (T1-3) carrying truncated CRMP2 were constructed as schematically indicated and applied to CBA assay (Figure 3C). T1 contained 141 amino acids N-terminus of isoform 1, T2, 36 N-terminus of isoform 2, and T3, 536 C-terminus which shared by three CRMP2 isoforms. The results indicate that the 536 aa C-terminus of CRMP2 is necessary for the immunoreaction (Figure 3D).

We also checked the IgG subtype of anti-CRMP2 antibody in the patients. As shown in Figure 3E, only IgG4 mediated anti-CRMP2 immunoreaction. Same results obtained with P2’s serum (data not shown). The results suggest that anti-CRMP2 IgG4 recognize the C-terminus of CRMP2 in our AE patients.

### Clinical features of the two patients with anti-CRMP2 antibody

P1 patient, a young adult female, was admitted due to persistent dizziness, visual rotation, recurrent nausea and vomiting and fever for 8 days, and unstable walking and blurred speech for one day. Fever was mild and the highest body temperature was 38.5L. Physical examinations revealed signs of cerebellum including poetry-like language, nystagmus, and bilateral finger-nose test instability. Slight neck stiffness was noticed.

CSF examination revealed a normal opening pressure of 148 mmH_2_O with only slightly elevated white blood cell count of 38 cells/µl (90% monocytes). Screenings of paraneoplastic antigens and serological autoantibodies including anti-nuclear, anti-thyroid peroxidase and anti-thyroglobulin antibodies were all negative. Serum anti-*M*.*P*. IgM tests were positive twice in one week after admission, and weak positive two weeks later. Serum anti-Widal’s O test was positive with a titer of 1:80 on admission and conversed to be negative two weeks later. Next generation sequencing (Vision Medicals, Guangzhou, China) of CSF sample detected 64 unique reads against *M*.*P*.. CSF test was negative for AE antibodies, but was positive by TBA.

During hospitalization, the patient’s symptoms deteriorated with recurrent nausea and vomiting, also her mental status was changed to be apathy. Physical examination showed opsoclonus-myoclonus involving her head and both arms. She was diagnosed as intracranial *M*.*P*. infection and possible secondary immune-mediated encephalitis. Azithromycin, doxycycline, intravenous methylprednisone (MP, 40mg/day) and intravenous immunoglobin (IVIG, 0.4 g/kg/d) were prescribed. Levodopa and clonazepam were also given to control the myoclonus. With the treatment, the patient’s symptoms improved with mild dizziness and opsoclonus. Cranial MRI was repeated and the result was not remarkable with mild white matter degeneration (Figure 4A-F). She was discharged to a local hospital for rehabilitation. During the follow-up study, she recovered partially with dizziness and a modified Rankin Scale (mRS) score of 2 at 3 months.

**Figure 4.**
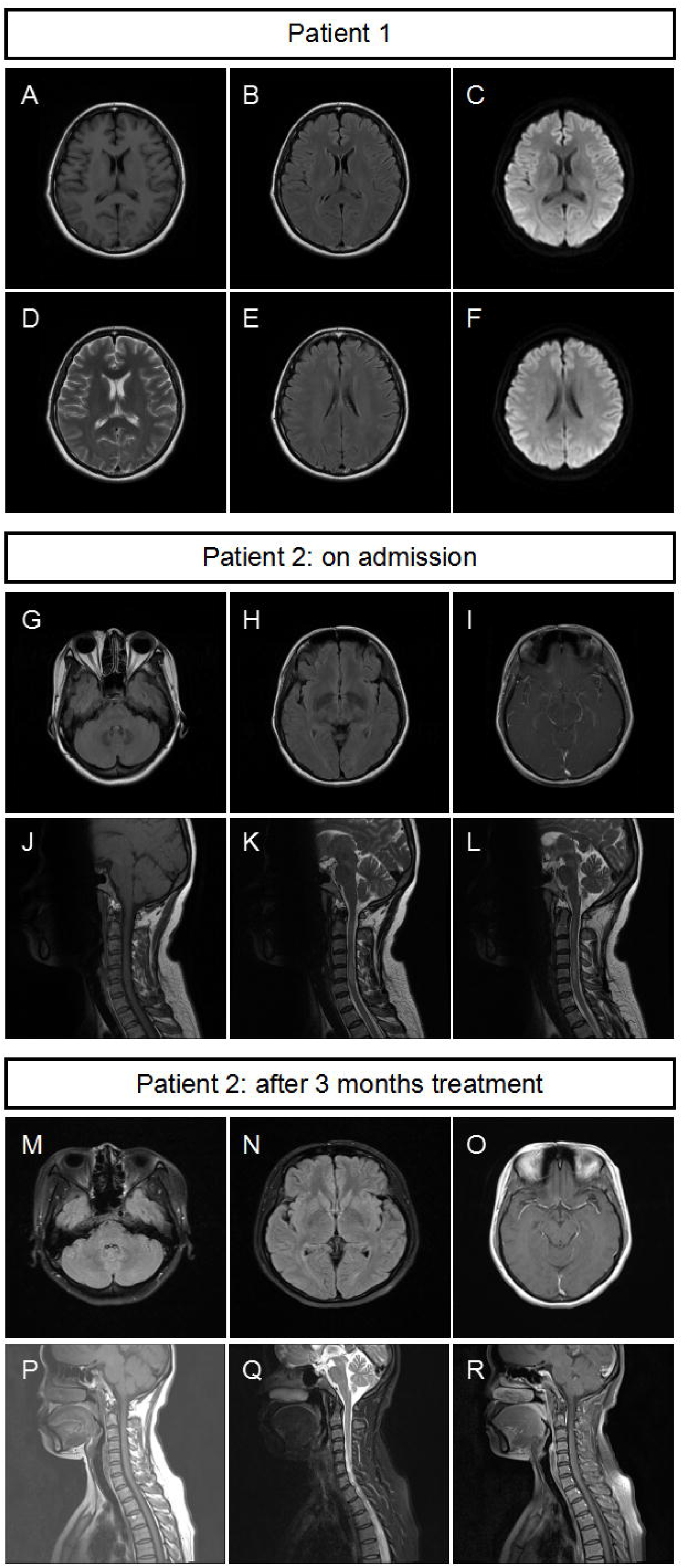
Magnetic resonance images of the patients. (A-F) Cranial MRI of patient 1 was not remarkable with mild whiter matter degeneration. (G-I) Cranial MRI of patient 2 showed multiple abnormal signals of white matters in bilateral cerebral hemisphere and brainstem. (J-L) Spine MRI of patient 2 showed long-segment spinal cord lesions from medulla to C6 segment. After 3 months of treatment, (M-O) the cranial MRI of patient 2 showed that the abnormal signals of the white matter and brainstem were reduced compared with G-I; (P-R) the cervical MRI showed that the original abnormal signals of the spinal cord had disappeared.

P2 patient, a young adult female, previously healthy, was admitted due to chest pain, right arm fatigue for 10 days and headache, nausea, vomiting, numbness in both legs for 6 days. Physical examination was not remarkable except for hyperalgesia in the left chest and abdomen, decreased needle tingling on the back of the left hand, and numbness on the feet. Chest CT and electrocardiogram (ECG) were not remarkable. Spine MRI showed long-segment spinal cord lesions from medulla to C6 segment (Figure 4J-L). Lumbar puncture revealed an opening pressure of 185 mmH_2_O, with moderately elevated WBC of 240 cells/µl (90% monocytes), decreased glucose of 2.64 mmol/l, normal chloride level of 123 mmol/l, and elevated protein level of 0.80 g/l. CSF pathogen screenings for tuberculosis (TB) culture, TB-SPOT, X-pert, bacteria and fungi cultures were all negative. Antibodies including autoimmune encephalitis panels, anti-aquaporin 4 (AQP4), anti-myelin oligodendrocyte glycoprotein (MOG) were all negative. Cranial MRI with contrast revealed multiple patchy abnormal signals in the white matter area of the bilateral cerebral hemispheres and brain stem (Figure 4G-I). TBA was negative for CSF, and positive for serum.

Possible tuberculous encephalomyelitis and acute demyelinating encephalomyelitis (ADEM) were considered. The patient was given anti-TB regimen (Rifampicin 0.45 g qd, Isoniazid 0.3 g qd, Pyrazinamide 0.5 g tid, Ethambutol 0.75 g qd) and intravenous MP pulse therapy (1 g/day for 5 days, and tapered to oral 48 mg/d). With the treatment, the patient’s chest pain and numbness were improved gradually. Lumber puncture was repeated 20 days later and revealed that the WBC decreased dramatically to 20 cells/µl, the glucose and protein returned to normal. She was followed up regularly in our clinic and the steroids was tapered. Anti-TB therapy was discontinued after 6 month’s treatment. Repeated cranial MRI revealed that the abnormal signals in the white matter area of the bilateral cerebral hemispheres and brain stem decreased dramatically (Figure 4M-O). Cervical MRI revealed that the abnormal signals of her spinal cord disappeared (Figure 4P-R).

The clinical features of the two patients were summarized in Table 1.

**Table 1.**
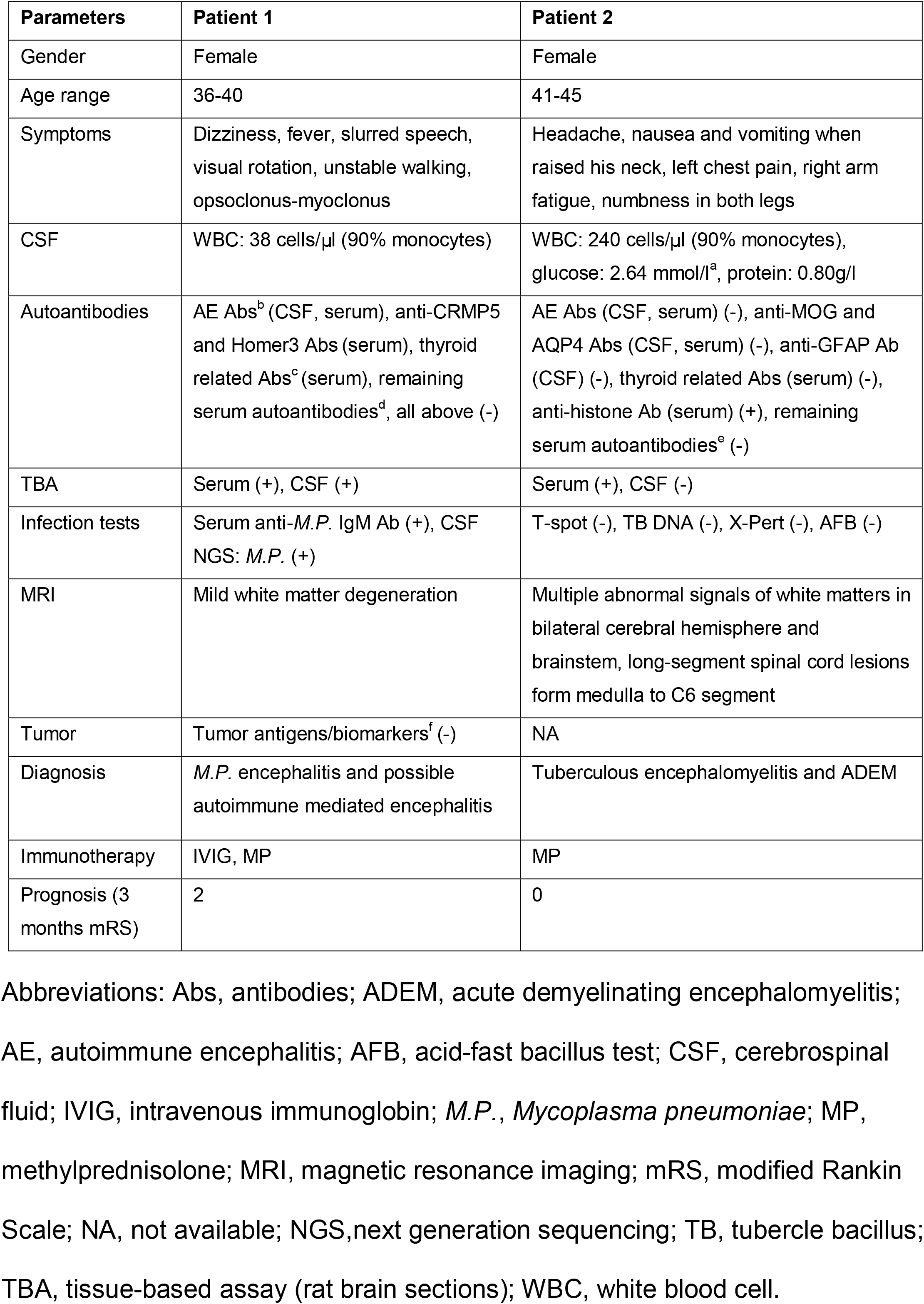
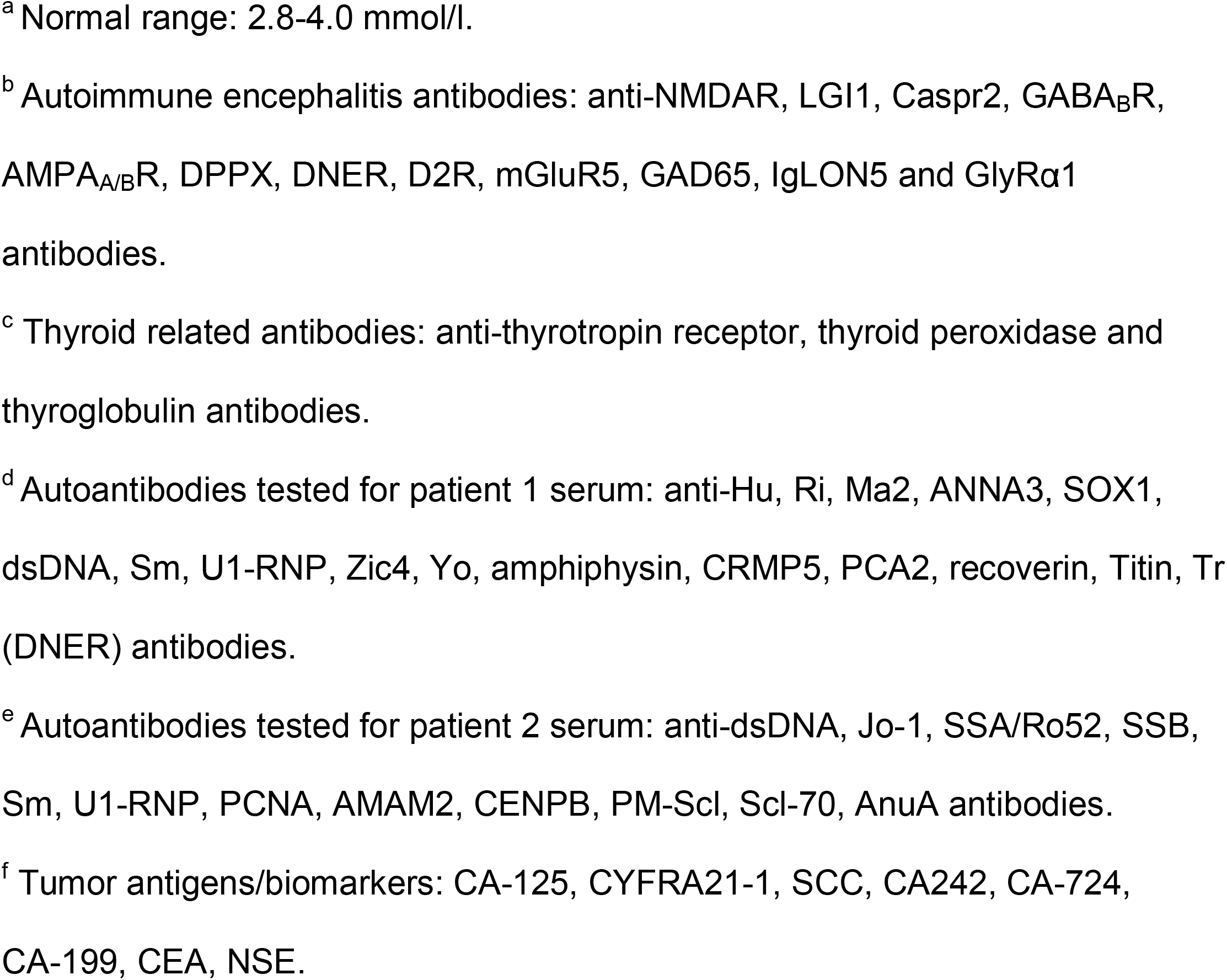
Clinical Features of the Patients with anti-CRMP2 antibody.

| Parameters | Patient 1 | Patient 2 |
| --- | --- | --- |
| Gender | Female | Female |
| Age range | 36-40 | 41-45 |
| Symptoms | Dizziness, fever, slurred speech, visual rotation, unstable walking, opsoclonus-myoclonus | Headache, nausea and vomiting when raised his neck, left chest pain, right arm fatigue, numbness in both legs |
| CSF | WBC: 38 cells/μl (90% monocytes) | WBC: 240 cells/μl (90% monocytes), glucose: 2.64 mmol/l<sup>a</sup>, protein: 0.80g/l |
| Autoantibodies | AE Abs<sup>b</sup> (CSF, serum), anti-CRMP5 and Homer3 Abs (serum), thyroid related Abs<sup>c</sup> (serum), remaining serum autoantibodies<sup>d</sup>, all above (-) | AE Abs (CSF, serum) (-), anti-MOG and AQP4 Abs (CSF, serum) (-), anti-GFAP Ab (CSF) (-), thyroid related Abs (serum) (-), anti-histone Ab (serum) (+), remaining serum autoantibodies<sup>e</sup> (-) |
| TBA | Serum (+), CSF (+) | Serum (+), CSF (-) |
| Infection tests | Serum anti-*M.P.* IgM Ab (+), CSF NGS: *M.P.* (+) | T-spot (-), TB DNA (-), X-Pert (-), AFB (-) |
| MRI | Mild white matter degeneration | Multiple abnormal signals of white matters in bilateral cerebral hemisphere and brainstem, long-segment spinal cord lesions form medulla to C6 segment |
| Tumor | Tumor antigens/biomarkers<sup>f</sup> (-) | NA |
| Diagnosis | *M.P.* encephalitis and possible autoimmune mediated encephalitis | Tuberculous encephalomyelitis and ADEM |
| Immunotherapy | IVIg, MP | MP |
| Prognosis (3 months mRS) | 2 | 0 |

Abbreviations: Abs, antibodies; ADEM, acute demyelinating encephalomyelitis; AE, autoimmune encephalitis; AFB, acid-fast bacillus test; CSF, cerebrospinal fluid; IVIG, intravenous immunoglobulin; *M.P.*, *Mycoplasma pneumoniae*; MP, methylprednisolone; MRI, magnetic resonance imaging; mRS, modified Rankin Scale; NA, not available; NGS, next generation sequencing; TB, tubercle bacillus; TBA, tissue-based assay (rat brain sections); WBC, white blood cell.
<sup>a</sup> Normal range: 2.8-4.0 mmol/l.
<sup>b</sup> Autoimmune encephalitis antibodies: anti-NMDAR, LGI1, Caspr2, GABA<sub>B</sub>R, AMPA<sub>A/B</sub>R, DPPX, DNER, D2R, mGluR5, GAD65, IgLON5 and GlyR $\alpha$ 1 antibodies.
<sup>c</sup> Thyroid related antibodies: anti-thyrotropin receptor, thyroid peroxidase and thyroglobulin antibodies.
<sup>d</sup> Autoantibodies tested for patient 1 serum: anti-Hu, Ri, Ma2, ANNA3, SOX1, dsDNA, Sm, U1-RNP, Zic4, Yo, amphiphysin, CRMP5, PCA2, recoverin, Titin, Tr (DNER) antibodies.
<sup>e</sup> Autoantibodies tested for patient 2 serum: anti-dsDNA, Jo-1, SSA/Ro52, SSB, Sm, U1-RNP, PCNA, AMAM2, CENPB, PM-Scl, Scl-70, AnuA antibodies.
<sup>f</sup> Tumor antigens/biomarkers: CA-125, CYFRA21-1, SCC, CA242, CA-724, CA-199, CEA, NSE.

## DISCUSSION

In this study, we identified IgG4-autoantibody specifically against CRMP2 in the sera and CSF samples of two AE patients with acute disease onset. Two patients had various symptoms. P1 presented with fever, nausea, vomiting, apathy, opsoclonus-myoclonus of head and arm, cerebellar signs including dizziness, slurred speech, and unstable walking. P2 had headache, nausea, vomiting, chest pain, right arm fatigue numbness in both legs. Both patients showed increased number of cells in CSF, and P2 had low glucose and increased level of protein. P1’s CSF had unique reads against *M*.*P*.. MRI images showed abnormal signals in the white matter in the two patients and long segment lesions on the spinal cord in P2. P1 was first diagnosed as *M*.*P*. infection. However, treatment with antibiotics only did not improve the symptoms. TBA immunostaining strongly indicated a secondary AE. Indeed, in additional of immunotherapy achieved good improvement at discharge and had 2 score of mRS at 3 months. P2 was diagnosis as possible TB & ADEM. 20 days treatment with anti-TB and immunotherapy reached good outcome for the patient at discharge and had 0 score of mRS at 3 months. Based on the study, we may correct the diagnosis as possible TB and autoimmune encephalomyelitis. No neoplasm was found in these patients currently and will be continuously follow up in the future. Our study provide evidence for the diagnosis of AE, expanding anti-CRMP2 antibody to the AE spectrum and suggesting to test anti-CRMP2 antibody in suspected AE patients.

CRMPs highly express during embryonic development and decreases significantly after birth, while each CRMP has a different subcellular localization ^22^. In adult mouse brain, CRMP2 is expressed in the soma, axons, and dendrites of neurons from the cortex, hippocampus, cerebellum, immature and mature oligodendrocytes ^23, 24^. It is thought that antibodies against intracellular antigen mediate T cell immunoreaction and impair cell function in the related area, while the antibodies themselves will not cause a pathogenic antibody-antigen interaction ^25-27^. Therefore, symptoms of the two patients are consistent with the subcellular distribution of CRMP2 in the brain, or probably in the spinal region. Indeed, sera /and CSF from the two patients with anti-CRMP2 antibody had robust staining of the cortex, CA3 region of hippocampus and Purkinje cells of cerebellum.

*M.P*. encephalitis is one of the most severe extra-respiratory complications, which accounts for 5-30% of all of the reported encephalitis cases^28^. The latency period between the onset of respiratory symptoms and the development of neurological abnormalities varies. In some cases, there was not a previous, clinically evident respiratory episode at all ^29^. P1 had definite intracranial infection of *M.P*. and early onset of encephalitis evidenced by the presence of IgM antibody against mycoplasma in serum and presence of *M.P*. DNA in CSF. Presence of antibody to CRMP2 suggests that P1 had an immediately immune-mediated CNS inflammation, which was also evidenced by the solely treatment with antibiotics deteriorated the patient’s symptoms. However, additional of IVIG and MP treatment improved her symptoms gradually. We considered that the symptoms of the patient were more attributed to the secondary inflammation caused by the *M.P*. infection. Anti-ganglioside and galactocerebroside antibodies have been found in association with mycoplasma neurological disorders. Thus, these studies and ours provide direct evidence that infection of *M.P*. cause secondary immune-mediated inflammation ^30^.

Infections by viral and bacterial pathogens are suspected to initiate a broad range of neurological inflammatory disorders particularly AE ^31^. Herpes simplex virus (HSV) has reported to trigger some types of AE frequently more than others, including anti-NMDAR, GABA_A_R, mGluR5, Neure-xin3a, and D2R antibodies. Among of them, near 64% AE patients post-HSV had anti-NMDAR antibody in their serum or CSF, while the remaining had antibodies unidentified. TB was associated with a lot of autoimmune disorders^32, 33^. Whether TB is a trigger of anti-CRMP2 encephalitis in P2 patient, more studies are warranted.

Each CRMP has distinct neuronal function. *Crmp2* gene knockout mouse shows that lack of CRMP2 leads to defects in axon guidance, axon pruning in hippocampus and visual cortex and altered dendritic spine remodeling^34^. The C-terminal region of CRMP2 involves in the post-translational modifications including phosphorylation, SUMOylation, oxidation, and O-GlcNAcylation. Several critical phosphorylation sites locate in the C-terminus^35-37^. The phosphorylated CRMP2 inhibits its interaction with tubulin heterodimers and augments interaction with NaV1.7^38^. CRMP2 binds to CaV2.2 and increases Ca^2+^ level on the cell surface, interacts with several receptors including NMDARs, Kainate, and NCX (Na^+^/Ca^2+^ exchanger 3) to regulate Ca^2+^ and Na^+^ neuronal homeostasis^39-40^. Although immunoreaction of anti-CRMP2 in related neurons is supposed not a direct causative reason, it might lead to dysregulation of other receptors. Epitope studies of CRMP2 indicated that C-terminus is responsive for the antibody binding, implying that antigen-antibody reaction may affect the function of CRMP2. The pathogenic properties of CRMP2 antibody should be clarified by more studies.

The present study consists small samples data from a single clinic center, thus, the detail clinical features and information of immunotherapy of anti-CRMP2 AE will be obtained in a large cohort study in the future. Our study characterized anti-CRMP2 antibody in two AE patients, who responded well to immunotherapy. We suggest to test anti-CRMP2 antibody in suggested AE patients.

## Supporting information

Table S1 in the supplement

## Data Availability

Data are available upon reasonable request. Anonymised data used in the present study may be available upon reasonable request to the corresponding author Y.F.H..

## ARTICAL INFORMATION

### Contributors

Y.F.H., S.Y.P., Y.M.W. and K.B.X. designed the study and drafted the manuscript; D.M.W. and S.N.W. took care of the index patient and responsible for the clinical data preparation; Y.H. and G.H.L. did most experiments; Y.P. provided cultured cells and neurons; H.S.J. and Y.P. were responsible for the other anti-CRMP2 antibody positive patients; F.L.X. prepared mouse and rat brain sections; Y.H.H. took confocal images; Q.Q.W. did Western blotting; all the authors reviewed the manuscript.

### Funding

This work was supported by the National Natural Science Foundation of China (81771225 to Y.F.H., 82071484 to Y.M.W.), Guangdong Provincial Scientific and Technologic Progression Fund (2016A020215182 to Y.F.H.), Natural Science Foundation of Guangdong Province (2019A1515011760 to Y.M.W.), President Foundation of Nanfang Hospital (2019B007 to D.M.W., 2020B008 to K.B.X.), Medical Science and Technology Foundation of Guangdong Province (A2021151 to K.B.X.).

### Patient consent for publication

The two patients with anti-CRMP2 antibody and clinical features presented signed the consents for publication.

