## Supplementary material for "Characterization of A Novel Autoimmune Encephalitis Associated Antibody against CRMP2": Table S1 in the supplement

**Table S1** LC-MS/MS identified CRMP2 peptides.

| **Peptide sequence** | **Expected mass** | **Calculated mass** | **Mass deviation** | **Score** | **Expected score^b^** |
| --- | --- | --- | --- | --- | --- |
| R.IVAPPGGR.A^a^ | 765.4508 | 765.4497 | 0.0011 | 13.96 | 0.44 |
| K.SAAEVIAQAR.K | 1014.547 | 1014.5458 | 0.0013 | 38.75 | 0.0036 |
| K.SAAEVIAQAR.K | 1014.5472 | 1014.5458 | 0.0014 | 26.69 | 0.057 |
| R.GSPLVVISQGK.I | 1083.6296 | 1083.6288 | 0.0008 | 7.92 | 2.4 |
| R.GSPLVVISQGK.I | 1083.6307 | 1083.6288 | 0.0019 | 24.04 | 0.059 |
| R.KPFPDFVYK.R | 1139.602 | 1139.6015 | 0.0004 | 19.21 | 0.24 |
| R.KPFPDFVYK.R | 1139.6021 | 1139.6015 | 0.0006 | 25.56 | 0.057 |
| R.KPFPDFVYK.R | 1139.6027 | 1139.6015 | 0.0012 | 17.21 | 0.37 |
| R.KPFPDFVYK.R | 1139.6034 | 1139.6015 | 0.0018 | 7.51 | 3.5 |
| R.KPFPDFVYK.R | 1139.6034 | 1139.6015 | 0.0019 | 26.98 | 0.039 |
| K.GIQEEMEALVK.D | 1245.6292 | 1245.6275 | 0.0018 | 0.98 | 24 |
| K.GIQEEMEALVK.D | 1261.6219 | 1261.6224 | -0.0004 | 2.31 | 15 |
| R.MVIPGGIDVHTR.F | 1293.6879 | 1293.6864 | 0.0015 | 11.31 | 1.6 |
| K.QIGENLIVPGGVK.T^a^ | 1322.7553 | 1322.7558 | -0.0004 | 22.64 | 0.041 |
| K.QIGENLIVPGGVK.T^a^ | 1322.7561 | 1322.7558 | 0.0003 | 18.6 | 0.11 |
| R.GLYDGPVCEVSVTPK.T | 1619.7861 | 1619.7865 | -0.0004 | 10.8 | 1.9 |
| K.DHGVNSFLVYMAFK.D | 1642.7813 | 1642.7814 | 0 | 4.96 | 5.3 |
| K.DHGVNSFLVYMAFK.D | 1642.7827 | 1642.7814 | 0.0013 | 57.66 | 0.000029 |
| K.IVLEDGTLHVTEGSGR.Y | 1681.8663 | 1681.8635 | 0.0028 | 42.51 | 0.00094 |
| K.IVLEDGTLHVTEGSGR.Y | 1681.8673 | 1681.8635 | 0.0037 | 34.81 | 0.0056 |
| K.IVLEDGTLHVTEGSGR.Y | 1681.8698 | 1681.8635 | 0.0063 | 25.93 | 0.039 |
| K.MDENQFVAVTSTNAAK.V^a^ | 1740.7937 | 1740.7989 | -0.0052 | 6.17 | 3.4 |
| K.MDENQFVAVTSTNAAK.V^a^ | 1740.802 | 1740.7989 | 0.0031 | 1.09 | 12 |
| K.MDENQFVAVTSTNAAK.V^a^ | 1740.8092 | 1740.7989 | 0.0103 | 21.12 | 0.13 |
| K.DNFTLIPEGTNGTEER.M | 1791.83 | 1791.8275 | 0.0025 | 42.62 | 0.00098 |
| R.SITIANQTNCPLYVTK.V | 1821.9279 | 1821.9295 | -0.0015 | 46.93 | 0.0004 |
| K.THNSALEYNIFEGMECR.G | 2085.8886 | 2085.8884 | 0.0002 | 37.54 | 0.00085 |
| K.AVGKDNFTLIPEGTNGTEER.M | 2147.0524 | 2147.0495 | 0.003 | 8.27 | 2.9 |
| R.NLHQSGFSLSGAQIDDNIPR.R | 2168.067 | 2168.061 | 0.006 | 56.88 | 0.000035 |
| K.GTVVYGEPITASLGTDGSHYWSK.N | 2424.1666 | 2424.1598 | 0.0068 | 14.44 | 0.61 |

^a^ Not unique peptides for CRMP2.

^b^ Peptides with expected score≤0.05 were considered significant.
